# Diagnostic performance of Alzheimer’s disease blood biomarkers in a Brazilian cohort

**DOI:** 10.1101/2025.02.23.24319116

**Authors:** Wyllians Vendramini Borelli, Pamela C L Ferreira, Wagner Scheeren Brum, João Pedro Ferrari-Souza, Giovanna Carello-Collar, Maila Holz, Victoria Tizeli, Matheus Zschornack Strelow, Carolina Formoso, Marcia Lorena Fagundes Chaves, Andreia Rocha, Cristiano Schaffer Aguzzoli, Francieli Rohden, Débora G. Souza, Artur Francisco Schumacher Schuh, Guilherme Povala, Bruna Bellaver, Pedro Rosa-Neto, Raphael Machado Castilhos, Tharick A. Pascoal, Eduardo R. Zimmer

## Abstract

**Background:** Blood-based biomarkers (BBMs) have emerged as promising tools to enhance Alzheimer’s disease (AD) diagnosis. Despite two-thirds of dementia cases occurring in the Global South, research on BBMs has predominantly focused on populations from the Global North. This geographical disparity hinders our understanding of BBM performance in diverse populations. To address this, we evaluated the diagnostic properties of AD BBMs in a real-world memory clinic from Brazil, one of the largest countries in the Global South. We measured blood and cerebrospinal fluid (CSF) biomarkers - amyloid-β (Aβ)40, Aβ42, phosphorylated tau (p-tau) 217, neurofilament light (NfL) chain, and glial fibrillary acidic protein (GFAP) - in 59 individuals. Sample comprised 20 cognitively unimpaired (CU) individuals, 22 with AD dementia, and 17 with vascular dementia (VaD). We compared BBM levels across diagnostic groups and assessed their discriminative ability for AD. Notably, individuals with VaD and AD had lower educational levels (6.8±3.0) compared to CU individuals (61.4±6.6). Among the BBMs tested, plasma p-tau217 demonstrated the best performance, exhibiting high accuracy in differentiating CU from AD (AUC 0.96) and Aβ pathology (AUC 0.98). However, the ability of AD BBMs to distinguish between AD and VaD was lower than expected (AUC from 0.52 to 0.79), particularly when compared to studies from the Global North. Our findings highlight the potential utility of BBMs for AD diagnosis in real-world settings within the Global South. However, they also underscore the need for proper implementation and validation of these biomarkers within these populations to ensure accurate and reliable results.

## Introduction

Alzheimer’s disease (AD) is the most prevalent cause of dementia worldwide^1^. Over the years, numerous studies revealed significant discrepancies between clinical AD diagnosis and neuropathological identification of core AD biomarkers – amyloid-β (Aβ) plaques and neurofibrillary tangles of phosphorylated tau (p-tau)^2^. It is remarkable that secondary memory clinics in high-income countries (HICs) with ample resources estimate that up to one-third of individuals clinically diagnosed with AD did not exhibit neuropathological features of the disease at autopsy^3^. In Low- and Middle-Income Countries (LMIC), where two-thirds of all individuals with dementia live^4^, this scenario may be even worse. This clinic-biological mismatch highlights the limitations in AD diagnosis^5^, underscoring the need for biomarkers to improve the accuracy of AD diagnosis in living patients.

The later development of biomarkers capable of detecting Aβ and tau pathologies through positron emission tomography (PET) and cerebrospinal fluid (CSF) exams revolutionized the field^6^. Subsequent research diagnostic criteria adopted these biomarkers and redefined AD as a clinical-biological entity^7^. However, the high costs of PET scans and the relative invasiveness of lumbar punctures have substantially hindered their widespread use, particularly in underrepresented populations^8^, exacerbating the gap in dementia research between HICs and LMICs.

In this regard, blood-based biomarkers (BBMs) have emerged as a less invasive and more cost-effective alternative^9-11^. Recently developed AD BBMs have demonstrated high accuracy in identifying brain Aβ and tau pathologies throughout the AD continuum^12-15^. Among BBM candidates, plasma p-tau217 has consistently demonstrated high accuracy in identifying brain Aβ in AD^15, 16^. In both primary and secondary care, p-tau217 stands out as the most promising biomarker for clinical implementation^17, 18^ due to its diagnostic performance comparable to well-established CSF biomarkers^19, 20^.

Despite the progress, the diagnostic performance of novel BBMs has been extensively studied in individuals from the Global North (such as Europe and USA) but remains underexplored in individuals from the Global South. BBMs offer a unique opportunity to address the gap in biomarker-based AD diagnosis in Global South countries due to their scalability and ease of collection. Proper validation is necessary to ensure a safe and accurate clinical implementation of BBMs in LMICs. In this regard, this study aims at evaluating the diagnostic performance of AD BBMs using a real-world cohort from a Brazilian memory clinic, composed primarily of patients with low educational attainment.

## Methods

### Participants

This study included 59 Brazilian individuals from the Neurology Service in Hospital de Clínicas de Porto Alegre (HCPA): 20 cognitively unimpaired (CU), 17 with vascular dementia (VaD), and 22 with AD dementia. Participants were clinically evaluated with a detailed review of their medical history and an interview with the participant and their study partner. Neurological and neuropsychological assessments were performed, including the Mini-Mental State Examination (MMSE) and the Clinical Dementia Rating (CDR). Clinical diagnosis of VaD and AD dementia were conducted in a tertiary memory clinic by cognitive neurologists according to international guidelines ^8, 21^. CU individuals had no subjective complaint or objective impairment, and a Clinical Dementia Rating (CDR) score of 0. AD and VaD groups had a CDR score of 1, classified as mild dementia^22^. Participants with inadequately treated systemic conditions with neurological impacts were not included. This study followed the Standards for Reporting of Diagnostic Accuracy (STARD) guidelines. The local ethics committee approved this study (#5.844.578) and written informed consent was obtained for all participants or their legal representatives.

### Biomarker collection and quantification

Blood samples were drawn from VaD, AD, and CU individuals following commercial ethylenediaminetetraacetic acid (EDTA) tubes, in accordance with previous protocols^22^ and HCPA guidelines. The tubes were then centrifuged at 2,000g for 10 minutes. CSF samples were collected from VaD and AD patients by lumbar puncture under local anesthesia using the aseptic technique, as previously described^23^. We collected 10 mL with polypropylene syringes, which were rapidly centrifuged at 4°C for 10 minutes at 10,000 g. CSF and plasma samples were aliquoted, stored at -80°C, and transferred under strict temperature control to the University of Pittsburgh, where they were thawed for the first time. We analyzed samples using the single-molecule array (Simoa) platform (Quanterix®) with the following kits: p-tau181 (V2 Advantage assay, #103714), p-tau217 (ALZpath)^24^ and Aβ42, Aβ40, NfL, and GFAP (Neurology 4-Plex E assay, #103670). P-tau181 was analyzed in CSF only, whereas other biomarkers were in both CSF and plasma.

### Statistical Analysis

Demographic characteristics are reported as mean (±SD) unless otherwise stated. Associations between biomarkers were tested using Spearman’s correlation. Linear models assessed biomarker differences across groups of interest adjusting for age and sex, and pairwise between-group contrast were performed using Sidak correction for controlling family-wise error rate. We also computed the mean percent change between symptomatic versus asymptomatic groups. The area under the receiver operating characteristic (ROC) curve (AUC) values were computed for the following outcomes: I) diagnostic accuracy of BBMs in differentiating CU from both dementia groups (VaD and AD); II) diagnostic accuracy of CSF and plasma biomarkers in differentiating VaD from AD; and III) diagnostic accuracy of plasma biomarkers in differentiating high from low Aβ levels in the CSF. Given that no previous cutoff for CSF Aβ42/40 ratio was available for this cohort, we leveraged the notable bimodal distribution of this biomarker to derive an exploratory cutoff for Aβ abnormalities using Gaussian mixture modelling ^24^, with the derived cutoff ≤ 0.07 falling in the middle of the bimodal distribution, as previously done in other memory clinic cohorts (**Figure S1**)^25, 26^.

Individuals above the cutoff were classified as low Aβ and below as high Aβ pathology, respectively. Differences in area under the ROC curve were tested using DeLong’s test with the pROC package in R (**Table S1**). Statistical significance was determined as *p* < 0.05, two-tailed. Statistical analyses were performed in R (v4.2.2.).

## Results

### Sample characteristics

Sample characteristics are reported in **Table 1**. The mean (SD) age of all participants was 70.7 (9.64) and most individuals were male (71.1%). Importantly, the average years of education was 10.3 (6.86). CU participants presented lower age (p < 0.01) and higher education (p < 0.001) than VaD and AD groups, but similar sex distribution (p = 0.72). Dementia groups (VaD and AD) presented no significant differences in age, years of education, and sex. Correlations between plasma biomarkers and clinical information are presented in **Figure 1**. Plasma Aβ42/40 ratio, p-tau217, NfL, and GFAP levels were correlated with age and education, while MMSE and CDR-SB were correlated only with p-tau217, NfL, and GFAP. Correlations between CSF and plasma biomarkers can be seen in **Figure S2**.

**Table 1.** Demographic characteristics of the study participants. Continuous variables are presented as mean (SD). Abbreviations: CU: Cognitively Unimpaired. VaD: Vascular Dementia. AD: Alzheimer’s Disease. MMSE: Mini-Mental State Examination. CDR-SB: Clinical Dementia Rating – Sum of Boxes. Aβ: amyloid-β. GFAP: glial fibrillary acidic protein. NfL: neurofilament light. P-tau: phosphorylated tau (p-tau).

|  | **CU**<br>**(n = 20)** | **VaD**<br>**(n = 17)** | **AD**<br>**(n = 22)** |
| --- | --- | --- | --- |
| Male, n (%) | 14 (70.0) | 11 (64.7) | 17 (77.3) |
| **Age, years** | 61.4 (6.6) | 73.6 (8.3) | 77 (5.9) |
| **Education, years** | 17.3 (6.9) | 7.5 (3.1) | 6.2 (2.9) |
| **Race** |  |  |  |
| Non-White, n (%) | 3 (15) | 6 (35.3) | 3 (13.6) |
| White, n (%) | 17 (85) | 11 (64.7) | 19 (86.4) |
| **MMSE score** | 27.5 (1.7) | 17.9 (5.12) | 16.7 (4.8) |
| **CDR-SB score** | 0 (0) | 6.9 (2.5) | 6.9 (2.1) |
| **CSF biomarkers** |  |  |  |
| p-tau181 (pg/mL) | - | 276.1 (118.2) | 809.8 (541.2) |
| p-tau217 (pg/mL) | - | 18.4 (13.9) | 104.8 (88.1) |
| Aβ42/40 (pg/mL) | - | 0.08 (0.02) | 0.05 (0.02) |
| GFAP (pg/mL) | - | 18339.4 (7649.9) | 17935.7 (6816.4) |
| NfL (pg/mL) | - | 9161.2 (15160) | 2143.2 (1573.6) |
| **Plasma biomarkers** |  |  |  |
| p-tau217 (pg/mL) | 0.13 (0.09) | 0.31 (0.18) | 0.81 (0.63) |
| Aβ42/40 (pg/mL) | 0.06 (0.01) | 0.05 (0.01) | 0.05 (0.01) |
| GFAP (pg/mL) | 84.9 (46.6) | 148.9 (58.1) | 197.7 (98.5) |
| NfL (pg/mL) | 17.3 (15.8) | 67.9 (91.2) | 30.9 (14.7) |

**Figure 1.**
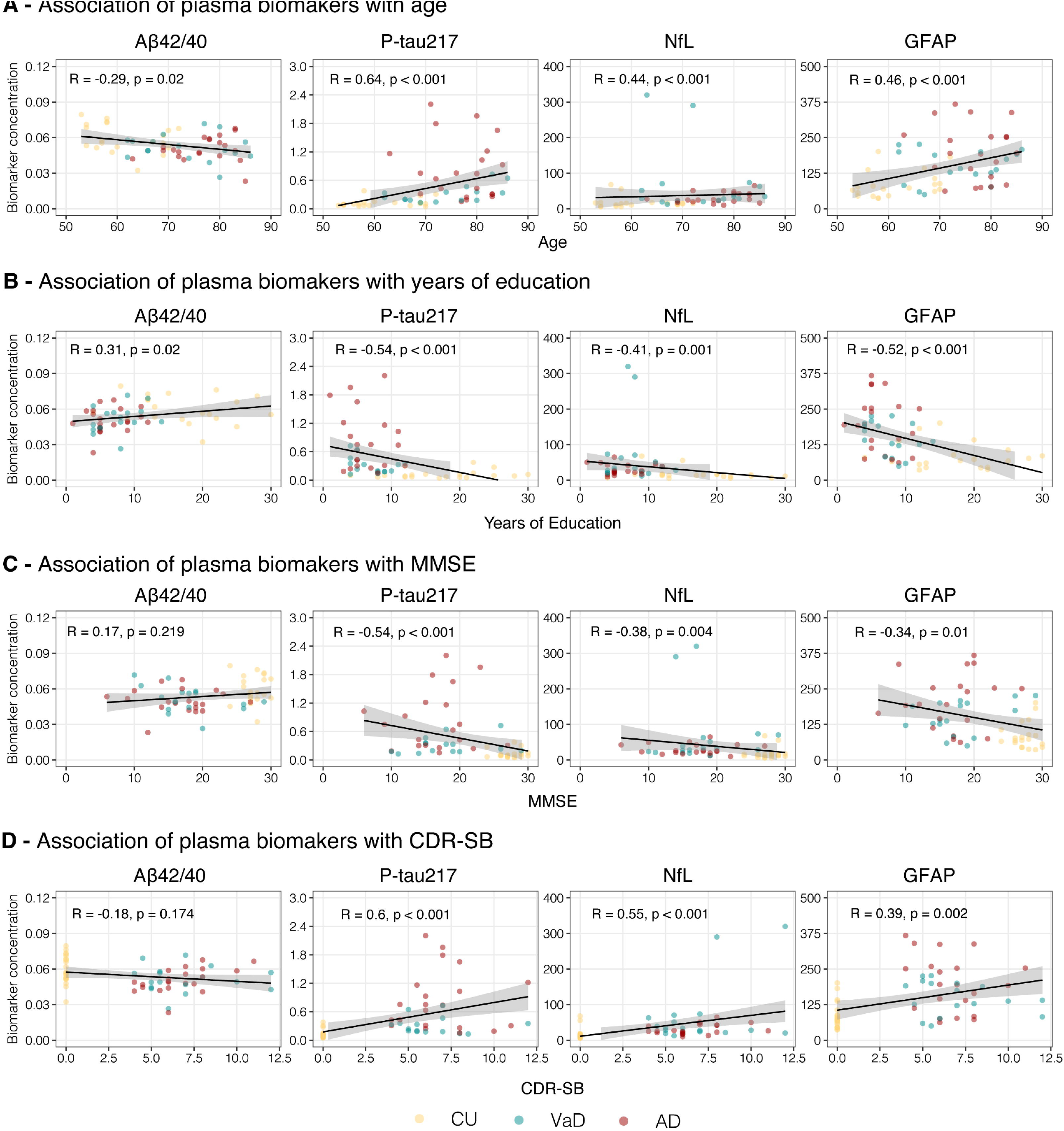
Correlation between plasma biomarkers and clinical variables. The panel shows the correlation between plasma biomarkers with (A) Age, (B) Education, (C) Mini-Mental State Examination, and (D) Clinical Dementia Rating – Sum of Boxes. Pearson’s coefficient (R) is demonstrated and its corresponding p-values.

### CSF biomarker levels across clinical diagnostic groups

**Figure 2A** demonstrates CSF biomarker levels in dementia participants (VaD and AD). We observed that CSF p-tau217, p-tau181, and Aβ42/40 ratio were significantly higher in AD compared to VaD (3.37, 1.63 and -0.27-fold-change, respectively; all p ≤ 0.05). CSF tau217/Aβ40 ratio was significantly higher in AD than VaD (2.54-fold-change, p < 0.01). CSF NfL and GFAP concentrations were not significantly different between AD and VaD (p = 0.14 and p = 0.47 respectively). Comparisons between groups were evaluated accounting by age, sex, and education.

**Figure 2.**
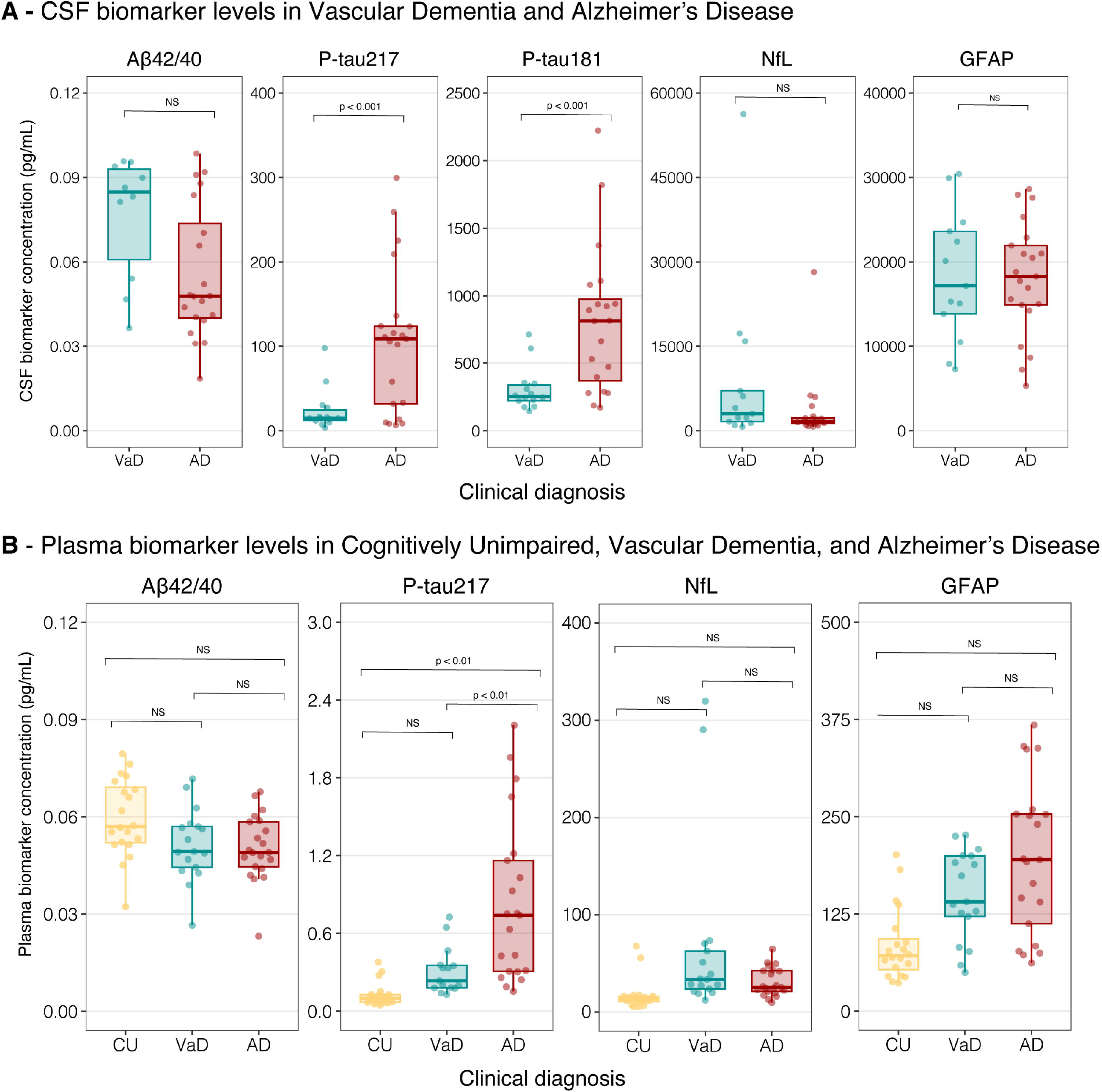
Plasma and CSF biomarkers levels distribution across clinical diagnosis. Panel A shows the CSF biomarker levels in VaD and AD individuals. Panel B shows the plasma biomarker levels across CU, VaD, and AD individuals. Comparison between groups were evaluated accounting by age, sex, and education. Abbreviations: CU: Cognitively Unimpaired. VaD: Vascular Dementia. AD: Alzheimer’s Disease.

### Blood-based biomarkers across clinical diagnostic groups

Plasma p-tau217 concentrations were significantly higher in AD individuals compared to CU (5.59, p ≤ 0.05) and VaD (1.45-fold change, p < 0.01) groups, while no significant differences were observed between CU and VaD (p = 0.88; **Figure 2B**). Plasma NfL, GFAP, and Aβ42/Aβ40 ratio concentrations did not differ significantly between AD, CU, and VaD individuals (all p > 0.05). Comparison between groups were evaluated accounting by age, sex, and education.

### Plasma biomarkers stratified by CSF Aβ burden

Plasma p-tau217 and Aβ42/40 ratio showed significant differences when the individuals were divided according to their CSF Aβ burden (p < 0.001; **Figure 3A-B**). In contrast, NfL and GFAP showed no significant differences between the groups (**Figure 3C-D**).

**Figure 3.**
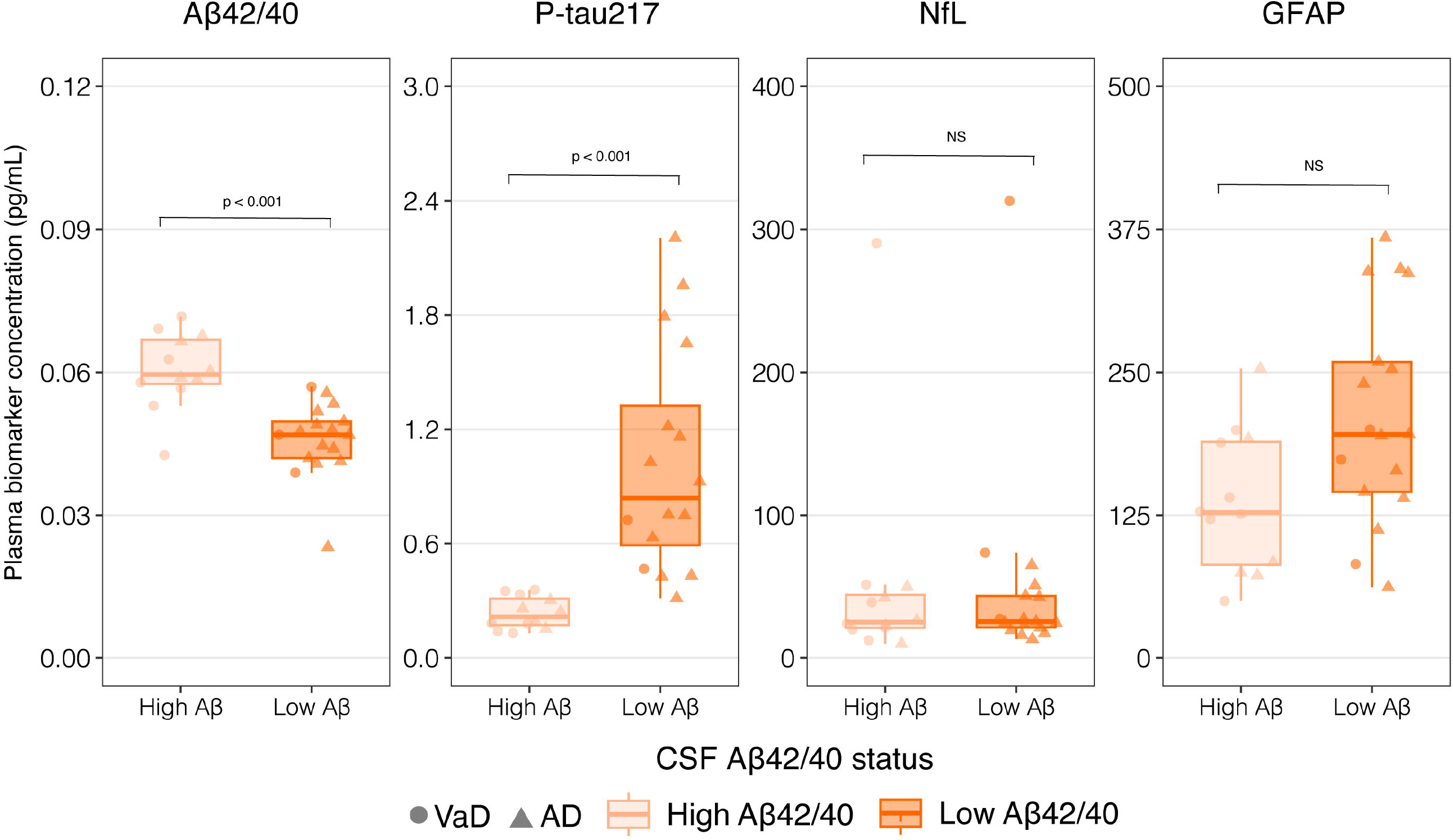
Plasma biomarker levels according to Aβ status. The Panel shows the plasma biomarker levels in VaD and AD individuals according to Aβ pathology burden. Comparison between groups were evaluated accounting by age, sex, and education. Abbreviations: CU: Cognitively Unimpaired. VaD: Vascular Dementia. AD: Alzheimer’s Disease.

### Diagnostic performance of biomarkers across clinical and biological diagnostic groups

Regarding the diagnostic performance of CSF biomarkers (**Figure 4A**), p-tau181 showed the highest diagnostic performance in discriminating AD and VaD when compared with GFAP. CSF p-tau181 showed the best performance discriminating VaD and AD (AUC 0.84 [95%CI 0.70 to 0.97]), but statistically similar to CSF p-tau217 (0.78 [95%CI 0.61 to 0.94]), Aβ42/40 ratio (0.73 [95%CI 0.54 to 0.92], NfL (0.69 [95%CI 0.48 to 0.89]), and GFAP (0.49 [95% CI 0.27 - 0.70]).

**Figure 4.**
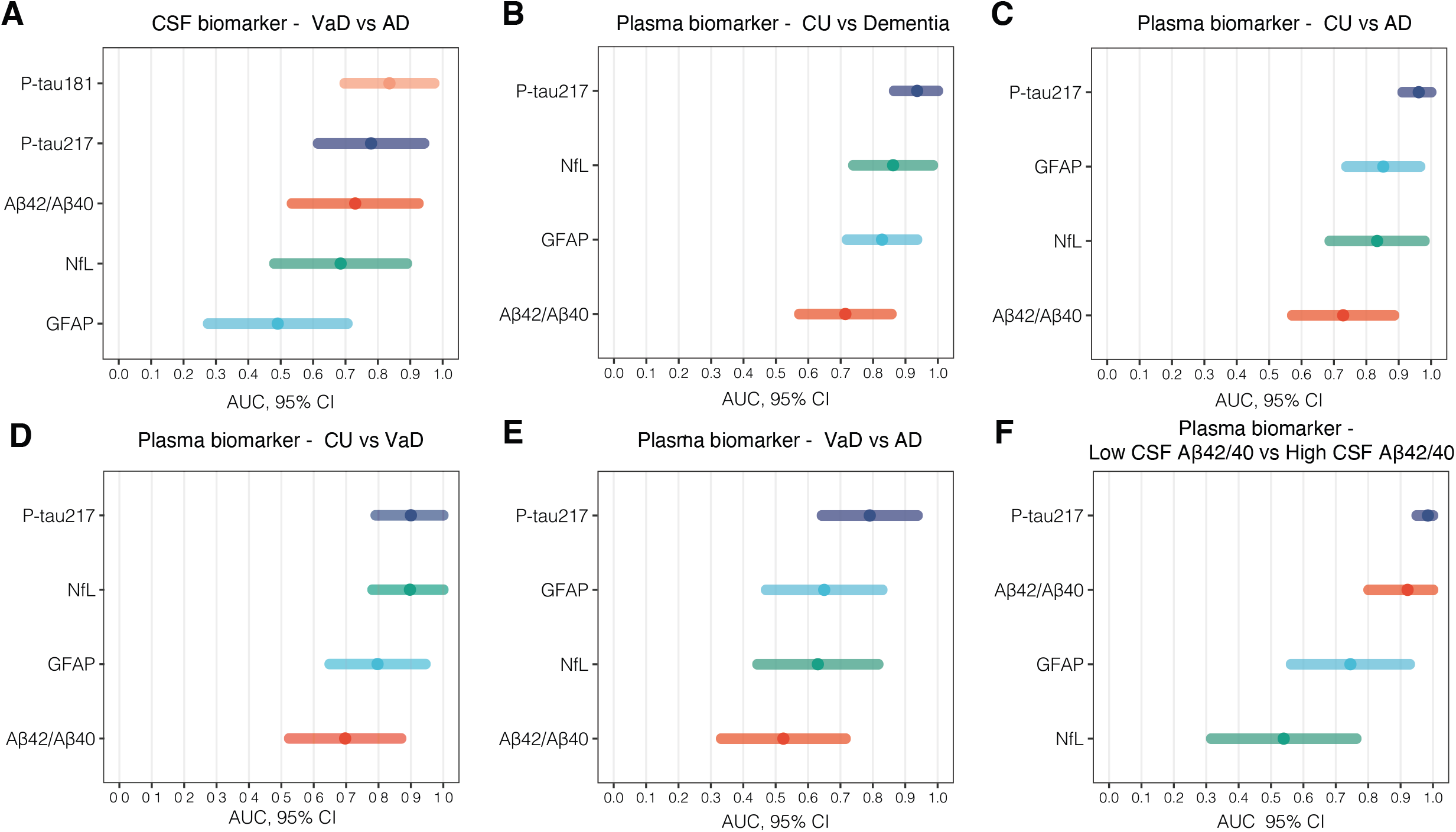
Diagnostic performance of biomarkers across clinical diagnosis. Panel A shows the AUC for CSF p-tau 181, p-tau 217, Aβ42/Aβ40, NfL, and GFAP levels in differentiating AD from VaD groups. Panel B shows the AUC for Plasma p-tau 217, NfL, GFAP, and Aβ42/Aβ40 in differentiating CU from dementia groups (AD and VaD combined). Panels C and D show the AUC in differentiating CU from AD and VaD, respectively. Panel E shows the AUC in differentiating AD from VaD. Panel F shows the AUC in differentiating low from high amyloid load, defined by CSF levels of Aβ42/Aβ40.

Plasma biomarkers differentiated CU from dementia (**Figure 4B**), with plasma p-tau217 exhibiting the best discriminatory performance (AUC = 0.94 [95%CI 0.86-1]), followed by NfL (AUC = 0.86 [95%CI 0.74-0.98]), GFAP (AUC = 0.83 [95%CI 0.72-0.94]), and the Aβ42/40 ratio (AUC = 0.72 [95%CI 0.57-0.86]). Notably, plasma p-tau217 significantly outperformed the Aβ42/40 ratio in differentiating CU from dementia (p = 0.006). Plasma p-tau217 differentiated CU from AD (**Figure 4C**), with high accuracy (AUC = 0.96 [95%CI 0.91-1]), followed by GFAP (0.85 [95%CI 0.74 to 0.97]), NfL (0.83 [95%CI 0.69 to 0.98]), and Aβ42/40 ratio (0.72 [95%CI 0.57 to 0.88]). Plasma p-tau217 significantly outperformed the Aβ42/40 ratio in differentiating CU from AD (p = 0.003), but did not differ significantly from NfL or GFAP. When comparing CU to VaD (**Figure 4D**), plasma p-tau 217 and NfL yielded higher performance to differentiate CU from VaD (plasma p-tau217 = AUC 0.90 [95%CI 0.78 - 1]; NfL = 0.90 [95%CI 0.78 to 1]), followed by GFAP (0.79 [95%CI 0.65 to 0.94]) and Aβ42/40 ratio (0.70 [95%CI 0.52 to 0.87]). In contrast, only plasma p-tau217 distinguished VaD from AD (**Figure 4E**) with moderate accuracy (AUC = 0.79 [95%CI 0.64-0.94]). **Figure 4F** showed that plasma p-tau217 accurately identified Aβ pathology (AUC 0.98 [95%CI 0.95 to 1]), together with plasma Aβ42/40 (AUC 0.92 [95%CI 0.8 to 1]). Plasma p-tau217 significantly outperformed NfL and GFAP in identifying Aβ pathology (p < 0.01 and p = 0.03, respectively).

## Discussion

We demonstrated the potential utility of BBMs for AD diagnosis in a Global-South real-world memory clinic-based cohort from Brazil. To the best of our knowledge, this is the first study to explore the diagnostic performance of novel ultrasensitive AD BBMs in a population of the Global South. Our findings demonstrate a high accuracy of BBMs in identifying abnormal Aβ pathology and diagnosing VaD and AD individuals, but a modest accuracy in distinguishing VaD from AD individuals. Moreover, BBMs presented similar diagnostic performance compared to CSF biomarkers (AUCs > 0.90 in some cases), showing the potential clinical utility of BBMs in resource-limited settings.

Notably, our cohort of individuals with AD and VaD had significantly lower levels of education (mean 6.79±3 years) compared to participants from HICs included in previous studies of plasma biomarkers (mean 15.3±3.6 years)^24 10^. This disparity is particularly relevant given that older adults with dementia in LMICs often exhibit low educational attainment, a characteristic that our sample reflects^27^. It is essential to consider the impact of educational attainment on cognitive reserve^28^, as it serves as a major proxy for neuroprotection against AD pathology^29^. Individuals with higher education tend to have greater cognitive reserves, which can provide substantial protection against AD-related cognitive decline. Considering this, it is crucial to investigate the diagnostic performance of BBMs in samples with low educational attainment and compare them to those with high educational backgrounds.

Our analysis revealed that plasma p-tau217 exhibited the highest diagnostic performance in accurately identifying CSF Aβ pathology, followed closely by the plasma Aβ42/40 ratio. In contrast, plasma GFAP and NfL showed lower diagnostic performance in detecting Aβ load, which is consistent with previous studies ^14, 30^ and their biological signifficance^31, 32^. Our findings align with a growing body of literature that supports the potential of plasma p-tau217 levels in identifying Aβ pathology in the brain^13, 15, 33, 34^. By demonstrating that plasma p-tau217 achieves sufficient accuracy in detecting Aβ pathology in this population, patients with early AD stages may also benefit from a biomarker-supported diagnosis. Notably, plasma Aβ42/40 has yielded inconsistent accuracy performances in previous studies^35^, but our results show that the high performance of the plasma Aβ42/40 ratio may be attributed to biological variations such as blood-brain barrier breakdown^36^ due to allostatic overload, which is more prevalent in individuals living in the Global South^37^.

The diagnostic performance of plasma biomarkers in differentiating between dementia due to AD or VaD was moderate in this population. Plasma p-tau217 demonstrated the highest discriminative ability in distinguishing mild AD from VaD groups, but its AUC of 0.79 was lower than that reported by studies conducted in HICs, where AUC values exceeded 0.90^11, 37-39^. This may be due to the unique genetic, environmental, and socio-cultural factors of the Latin American populations^40^. Another potential explanation is the co-existence of amyloid pathology in VaD patients, as there are shared risk factors between these conditions^41^. Countries in the Global South face a higher burden of uncontrolled risk factors^42^, which directly contributes to the high allostatic load characteristic of these regions^43^. Collectively, these findings highlight the need for larger studies to evaluate the discriminative performance of plasma biomarkers in LMICs.

The clinical use of BBMs in Brazil and other countries of the Global South without thorough evaluation in real-world populations, including validation using gold-standard measurements, raises significant concerns. Notably, demand for commercially available blood tests has been increasing among healthcare professionals in Brazil in recent years, despite the fact that these tests have not yet been validated in the Brazilian population or approved as an “in vitro diagnostic product” by the Brazilian Health Regulatory Agency (ANVISA). To address this issue, BBMs should follow a structured roadmap for validation and implementation in these populations, as premature clinical use of these biomarkers may have adverse consequences for public health^44^. It is essential to consider that Brazil is currently implementing dementia plans^45^ and aims to establish a National Dementia Plan by 2025. The successful integration of BBMs has the potential to be a game-changer in secondary prevention, particularly given the unique context of the Brazilian Unified Health System (SUS), one of the largest government-run public healthcare systems worldwide, which provides free healthcare services to all citizens^46^.

The strengths of this study include the use of data collected from a population in Brazil with low educational attainment levels. However, our findings must be interpreted with caution, considering several limitations. Firstly, CU individuals had higher education and lower age levels than individuals with dementia. To mitigate this limitation, we controlled for age and education in the regression models. Secondly, only AD and VaD individuals underwent CSF analysis due to ethical considerations in our cohort. Furthermore, this study is subject to further limitations, including small sample size, limited availability of CSF samples from AD and VaD individuals only, disparities in age and educational attainment between groups, lack of amyloid and tau PET scans, and absence of APOE genotyping. Therefore, we acknowledge that these findings require replication in larger cohorts to confirm their validity.

In summary, our findings demonstrate that plasma AD biomarkers, particularly p-tau217, exhibit high diagnostic accuracy in identifying CSF Aβ pathology and cognitive impairment, as well as moderate diagnostic accuracy in distinguishing VaD from AD in a Brazilian memory center population. Our results have significant implications for public health strategies in LMICs, where access to CSF tests and PET scans is often limited or unaffordable for most individuals with dementia. The diagnostic properties of plasma biomarkers may offer a valuable alternative for improving dementia diagnosis and care in these regions.

## Data Availability

All data produced in the present study are available upon reasonable request to the authors

## Acknowledgements

We acknowledge all participants that voluntarily provided time and effort for this study.

This project was partly funded by FIPE/HCPA. WVB receives financial support by the Alzheimer’s Association (AACSFD-22-928689). PCLF is supported by the Alzheimer’s Association (AARFD-22-923814). BB is supported by the Alzheimer’s Association (grant No. AARFD-22-974627) and the National Institute of Aging (5 P01 AG025204-17). CSA is supported by the Global Brain Health Institute, Alzheimer’s Association, and Alzheimer’s Society (grant No. GBHI ALZ UK-23-971089), and Alzheimer’s Association (grant No. 24AACSF-1200375). DG is supported by the Alzheimer’s Association (AARFD-22-928702). GP is supported by the Alzheimer’s Association (24AARFD-1243899). PRN is supported by the Fonds de Recherche du Québec – Santé (FRQS; Chercheur Boursier, 2020-VICO-279314) and Colin J. Adair Charitable Foundation. TAP is supported by the NIA (5R01AG075336, 5R01AG073267). ERZ receives financial support from CNPq [312410/2018-2; 435642/2018-9; 312306/2021-0; 409066/2022-2], ARD/FAPERGS [21/2551-0000673-0], Alzheimer’s Association [AARGD-21-850670], CNPQ/FAPERGS/PRONEX [16/2551-0000475-7], the Brazilian National Institute of Science and Technology in Excitotoxicity and Neuroprotection [465671/2014-4], Instituto Serrapilheira [Serra-1912-31365], and Alzheimer’s Association and National Academy of Neuropsychology [ALZ-NAN-22-928381].

## Supplementary information is available at MP’s website

