## Supplementary material for "Diagnostic performance of Alzheimer’s disease blood biomarkers in a Brazilian cohort": Sup. Material

**Supplementary Figure** 1 – Histogram and density plot of CSF Aβ42/40 ratio distribution.

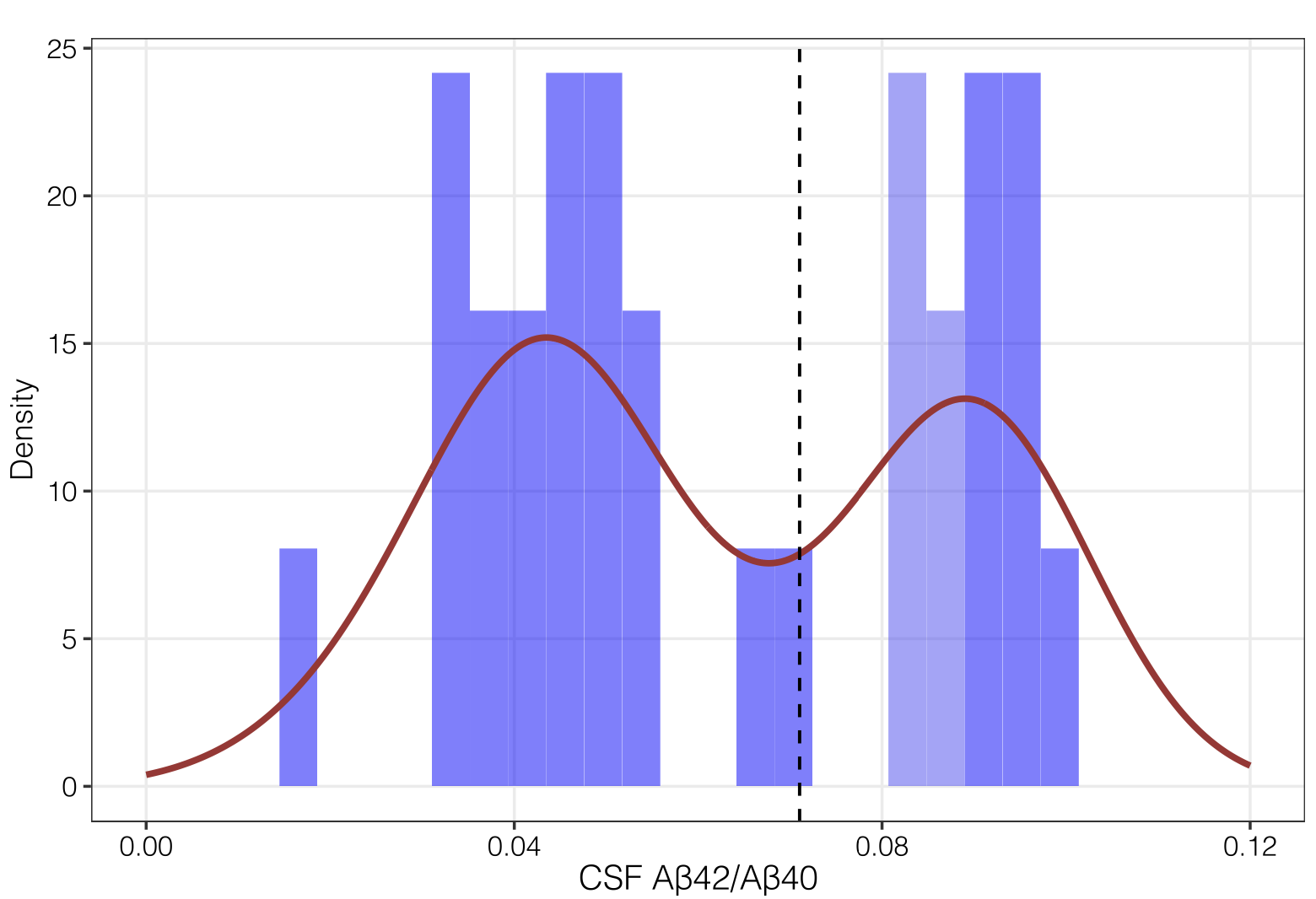

**Supplementary Figure 2** – Correlation between plasma and CSF biomarkers.

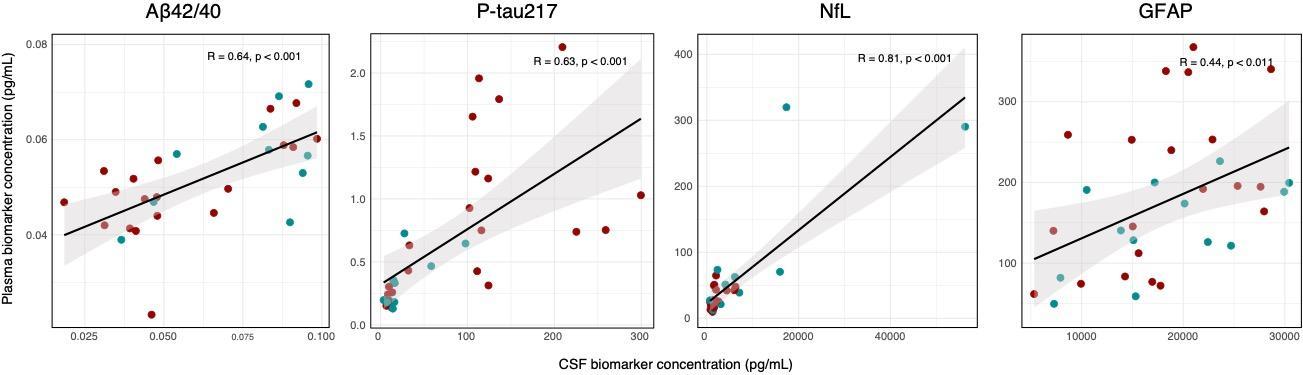

**Supplementary Table 1.** Fluid biomarkers mean (SD), mean fold-change, statistical tests, and effect sizes between low and high CSF Aβ pathology.

|  | **VaD** | **AD** | **Fold-change** | **Comparison t-value** | **p-value** | **Effect size** |
| --- | --- | --- | --- | --- | --- | --- |
| CSF Aβ42/Aβ40 | 0.076 (0.02) | 0.056 (0.02) | -0.26 | 2.37 | 0.03 | 0.90 |
| Plasma Aβ42/Aβ40 | 0.050 (0.01) | 0.051 (0.01) | 0.02 | 0.27 | 0.79 | 0.09 |
| CSF p-tau217/Aβ42 | 0.063 (0.08) | 0.414 (0.4) | 5.57 | -4.28 | 0.0002 | -1.21 |
| Plasma p-tau217/Aβ42 | 0.044 (0.02) | 0.156 (0.1) | 2.54 | -3.47 | 0.002 | -1.03 |
| CSF p-tau217 | 24.029 (25.1) | 104.987 (85.9) | 3.37 | -4.06 | 0.0004 | -1.18 |
| Plasma p-tau217 | 0.31 (0.2) | 0.83 (0.6) | 1.67 | -3.61 | 0.001 | -1.06 |
| CSF NfL | 9161.275 (±15160.686) | 3384.917 (±5894.143) | -0.63 | 1.31 | 0.21 | 0.56 |
| Plasma NfL | 67.9 (91.2) | 30.9 (14.7) | -0.54 | 1.65 | 0.11 | 0.60 |
| CSF GFAP | 18339.4 (7649.9) | 18079.7 (6676.4) | -0.01 | 0.1 | 0.92 | 0.04 |
| Plasma GFAP | 158.9 (58.1) | 197.7 (98.6) | 0.24 | -1.89 | 0.06 | -0.59 |

Note: p-tau biomarker means are reported in pg/ml. t-tests were carried out using log-transformed p-tau biomarker data. Effect sizes are reported as Cohen's d. Abbreviations: VaD: Vascular Dementia. AD: Alzheimer’s Disease.
